# Human monoclonal antibodies block the binding of SARS-CoV-2 spike protein to angiotensin converting enzyme 2 receptor

**DOI:** 10.1101/2020.04.06.20055475

**Authors:** Xiangyu Chen, Ren Li, Zhiwei Pan, Chunfang Qian, Yang Yang, Renrong You, Jing Zhao, Pinghuang Liu, Leiqiong Gao, Zhirong Li, Qizhao Huang, Lifan Xu, Jianfang Tang, Qin Tian, Wei Yao, Li Hu, Xiaofeng Yan, Xinyuan Zhou, Yuzhang Wu, Kai Deng, Zheng Zhang, Zhaohui Qian, Yaokai Chen, Lilin Ye

## Abstract

The severe acute respiratory syndrome coronavirus 2 (SARS-CoV-2) has caused a global pandemic of novel corona virus disease (COVID-19). To date, no prophylactic vaccines or approved therapeutic agents are available for preventing and treating this highly transmittable disease. Here we report two monoclonal antibodies (mAbs) cloned from memory B cells of patients recently recovered from COVID-19, and both mAbs specifically bind to the spike (S) protein of SARS-CoV-2, block the binding of receptor binding domain (RBD) of SARS-CoV-2 to human angiotensin converting enzyme 2 (hACE2), and effectively neutralize S protein-pseudotyped virus infection. These human mAbs hold the promise for the prevention and treatment of the ongoing pandemic of COVID-19.

## Main text

According to World Health Organization (WHO) newly updated situation report on March 18^th^, 2020, the corona virus disease 2019 (COVID-19) pandemic has confirmed 191,127 cases and claimed 7,807 deaths worldwide^1^. The etiological agent of COVID-19 has been identified as a novel coronavirus, the severe acute respiratory syndrome coronavirus 2 (SARS-CoV-2), belonging to *Sarbecovirus* subgenus (genus *Betacoronavirus*, family *Coronaviridae*) and showing 79.6% and 96.2% sequence identity in nucleotide to SARS-CoV and a bat coronavirus (BatCoV RaTG13), respectively^2-4^. Like SARS-CoV infection, a substantial fraction of COVID-19 patients exhibits severe respiratory symptoms and has to be hospitalized in intensive care unit (ICU)^5-8^. Although the mortality rate of COVID-19 is significantly lower than that of SARS-CoV infection, SARS-CoV-2 shows much higher human-to-human transmission rate, rapidly leading to a global pandemic declared by WHO on March 11^th^, 2020^9^.

Currently, there are no approved prophylactic vaccines or therapeutic drugs that are specific to COVID-19. Blocking monoclonal antibodies (mAbs), due to their extraordinary antigen specificity, are one of the best candidates for neutralizing virus infection^10, 11^. Therefore, identifying and cloning blocking mAbs that can specifically target surface viral proteins to block the viral entry to host cells is a very attractive approach for preventing and treating COVID-19, in particular when effective vaccines and therapeutics are unavailable in the outbreak of the COVID-19 pandemic. We then sought to identify and clone blocking mAbs from the memory B cell repertoire of recently recovered COVID-19 patients to prevent the entry of COVID-19 virus to the host cells.

Similar to SARS-CoV, SARS-CoV-2 also utilizes highly glycosylated homotrimeric spike (S) protein for receptor binding and virus entry^3, 12-15^. The S protein of SARS-CoV-2 consists of two subunits, S1 and S2. To engage host cell receptor human angiotensin-converting enzyme 2 (hACE2), shared by both SARS-CoV and SARS-CoV-2, S protein undergoes dramatic conformational changes to expose the RBD and key residues for receptor binding. S protein is metastable, and binding of RBD to hACE2 receptor likely leads to the shedding of S1 protein from S2 protein, thus promoting S2-mediated virus-host membrane fusion and virus entry^16-18^. Given the critical role of the RBD in initiating invasion of SARS-CoV-2 into host cells, it becomes a vulnerable target for neutralizing antibodies. Thus far, the human mAbs specifically target the SARS-CoV-2 RBD-hACE2 interaction have not been reported, and a monoclonal antibody targeting S1 made from immunized transgenic mice expressing human Ig variable heavy and light chains has been recently shown to neutralize both SARS-CoV-2 and SARS-CoV infection, but by an unknown mechanism that is independent of the blockade of RBD-hACE2 interaction^19^.

Prior to cloning SARS-CoV-2 RBD specific human mAbs, we first examined whether patients recently recovered from COVID-19 had mounted anti-SARS-CoV-2 S1 protein IgG antibodies in sera. Among 26 recovered COVID-19 patients, we found that the majority of these recruited patients were able to produce high titers of SARS-CoV-2 S1-specific IgG antibodies and only 3 patients mounted relatively lower anti-S1 IgG responses, by enzyme-linked immunosorbent assay (ELISA) (Fig. 1a). Consistently, we also found that SARS-CoV-2 RBD specific IgG antibodies were present in sera of all patients by ELISA (Fig. 1b). Next, we sought to investigate whether RBD-specific antibodies in patient serum can block the binding of SARS-CoV-2 RBD to hACE2. To this end, we set up an ELISA-based inhibition assay to examine the blocking function of these antibodies. We noted that there were only 3 out of 26 patients showed effective blockade of SARS-CoV-2 RBD binding to hACE2 (Fig. 1c). Taken together, these results suggested that while all recovered COVID-19 patients can generate anti-S1 and anti-RBD antibodies, there were only a small fraction of these antibodies can block the binding of RBD to hACE2 receptor. This observation may be explained by transient and dynamic perfusion conformational states of S protein that provide very limited window for the immunogenic epitopes of RBD exposure to specific B cells^20^.

**Figure 1.**
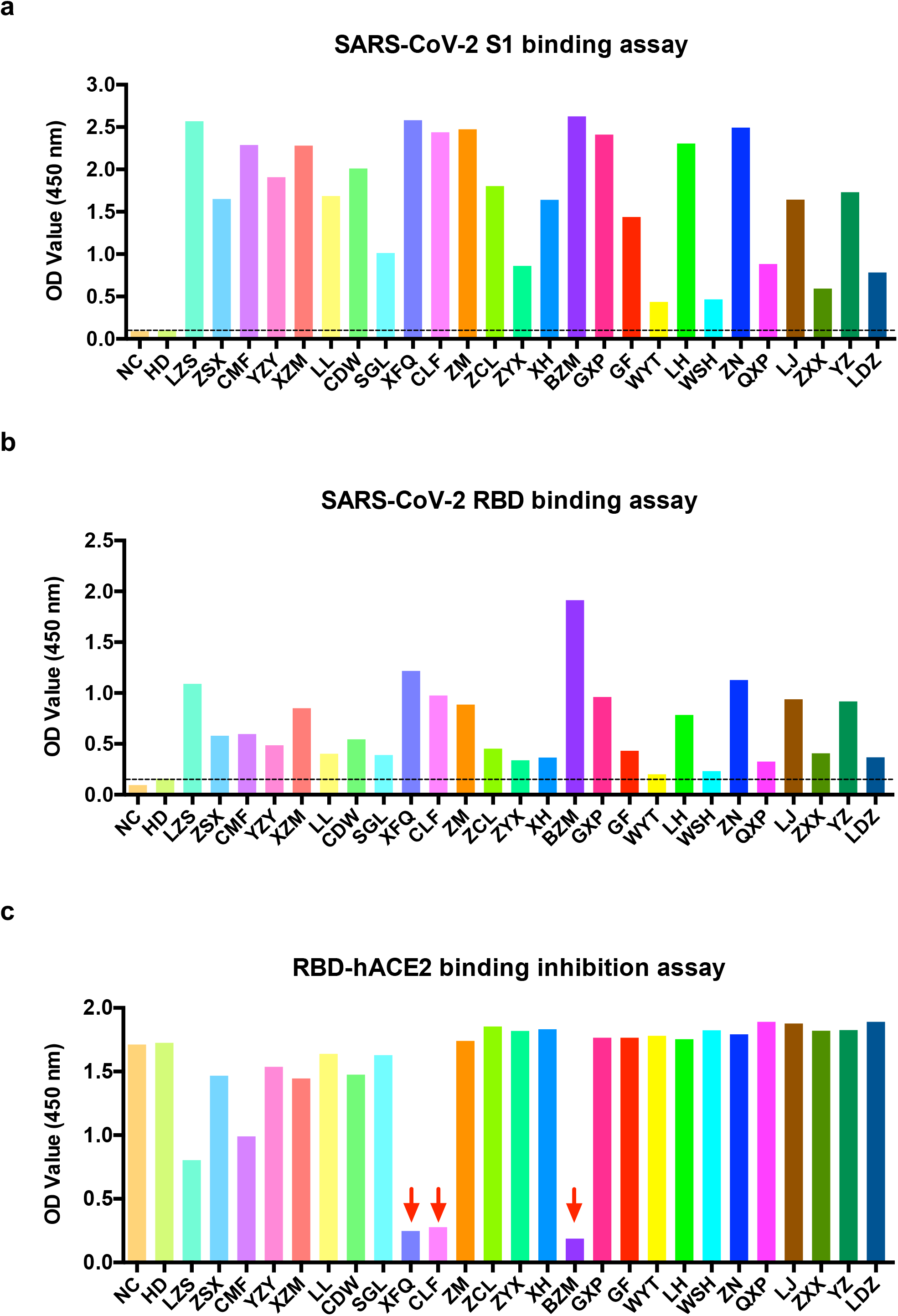
ELISA binding and blocking assays of COVID719 patient sera. **(a)** ELISA binding assay of COVID-19 patient sera to ELISA plate coating of SARS-CoV-2 S1 protein. **(b)** ELISA binding assay of COVID-19 patient sera to ELISA plate coating of SARS-CoV-2 RBD protein. **(c)** COVID-19 patient serum-mediated inhibition of the SARS-CoV-2 S1 protein binding to hACE2 protein by ELISA. NC, negative control. HD, healthy donor.

Next, we set out to clone human monoclonal antibodies using the blood samples from three COVID-19 recovered patients, of which their sera showed potent hACE2 receptor binding inhibition. To this end, we sorted each SARS-CoV-2 RBD specific, IgG class-switched memory B cell into a single well of the 96-well microplates. Subsequently, we used reverse transcription polymerase chain reaction (RT-PCR) to amplify IgG variable heavy chain (VH) and light chain (VL) from each single memory B cell. After cloning both VH and VL, we inserted both sequences into expression plasmids that encoding constant regions of human IgG1 heavy chain and light chain (Fig. 2a)^21^. We found that SARS-CoV-2 RBD specific, IgG-positive memory B cells only enriched in COVID-19 recovered patients, but not in healthy controls (Fig. 2b), suggesting the specificity of our sorting strategy. The representative RT-PCR and cloning of IgG1 heavy chains and light chains to expression plasmids were shown in Figure 2c and 2d. After antibody cloning, we acquired 3 pairs of IgG VHs and VLs inserted expression plasmids and the CDR3 sequences of heavy chains were shown in Figure 2e (analyzed with IMGT browser, http://www.imgt.org/IMGT_vquest/analysis#sequence1_alj).

**Figure 2.**
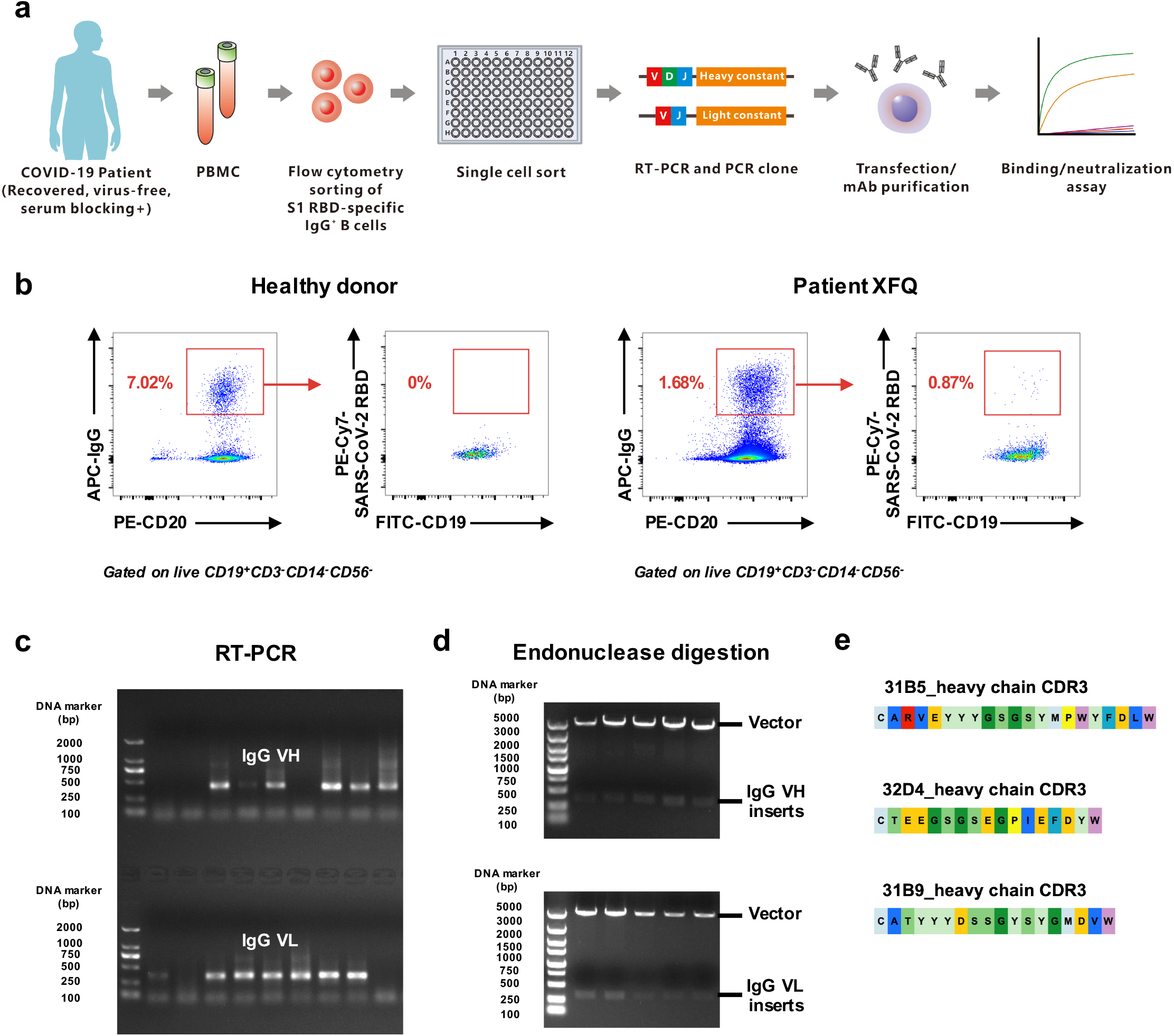
Cloning human mAbs from single SARS-Cov2 RBD-specific IgG^+^ memory B cells of three recovered patients whose sera potentially blocked RBD-hACE2 interaction. **(a)** An overall strategy of anti-SARS-CoV-2 RBD mAbs. **(b)** Flow cytometry analysis of SARS-CoV-2 RBD-specific IgG^+^ B cells in PBMCs of healthy donor and patient XFQ. **(c)** Heavy and light Ig-chain encoding segments of SARS-CoV-2 RBD-specific IgG^+^ B cells were amplified by single-cell RT-PCR. **(d)** Endonuclease digestion of IgG VH and VL inserts in the plasmids. **(e)** CDR3 sequences of heavy chains.

Finally, we expressed these paired plasmids encoding IgG VH and VL sequences and named these three mAbs as 311mab-31B5, 311mab-32D4 and 311mab-31B9, respectively. We first examined whether these human mAbs were able to bind to SARS-CoV-2 RBD protein by ELISA. The results showed that all three mAbs strongly and specifically bind to the RBD protein (Fig. 3a). Next, we tested whether these mAbs can block the interaction between SARS-CoV-2 RBD and hACE2. We found that both 311mab-31B5 and 311mab-32D4 could efficiently block SARS-CoV-2 RBD-hACE2 interaction (IC_50_=0.0332, and 0.0450 μg/ml, respectively), while 311mab-31B9 clone failed to inhibit such an interaction (Fig. 3b). The 31B5- and 32D4-mediated inhibition of RBD-hACE2 interaction was also evidenced by flow cytometry analysis (Fig. 3c-e). Furthermore, we determined the neutralization of these three mAbs using a SARS-CoV-2 S pseudotyped lentiviral particle^22^. In line with ELISA- and flow cytometry-based blockade results, both 311mab-31B5 and 311mab-32D4 effectively neutralized pseudovirus entry to host cells ectopically expressing hACE2 (IC_50_=0.0338, and 0.0698 μg/ml, respectively). As expected, 311mab-31B9 clone failed to show any neutralization activities (Fig. 3f).

**Figure 3.**
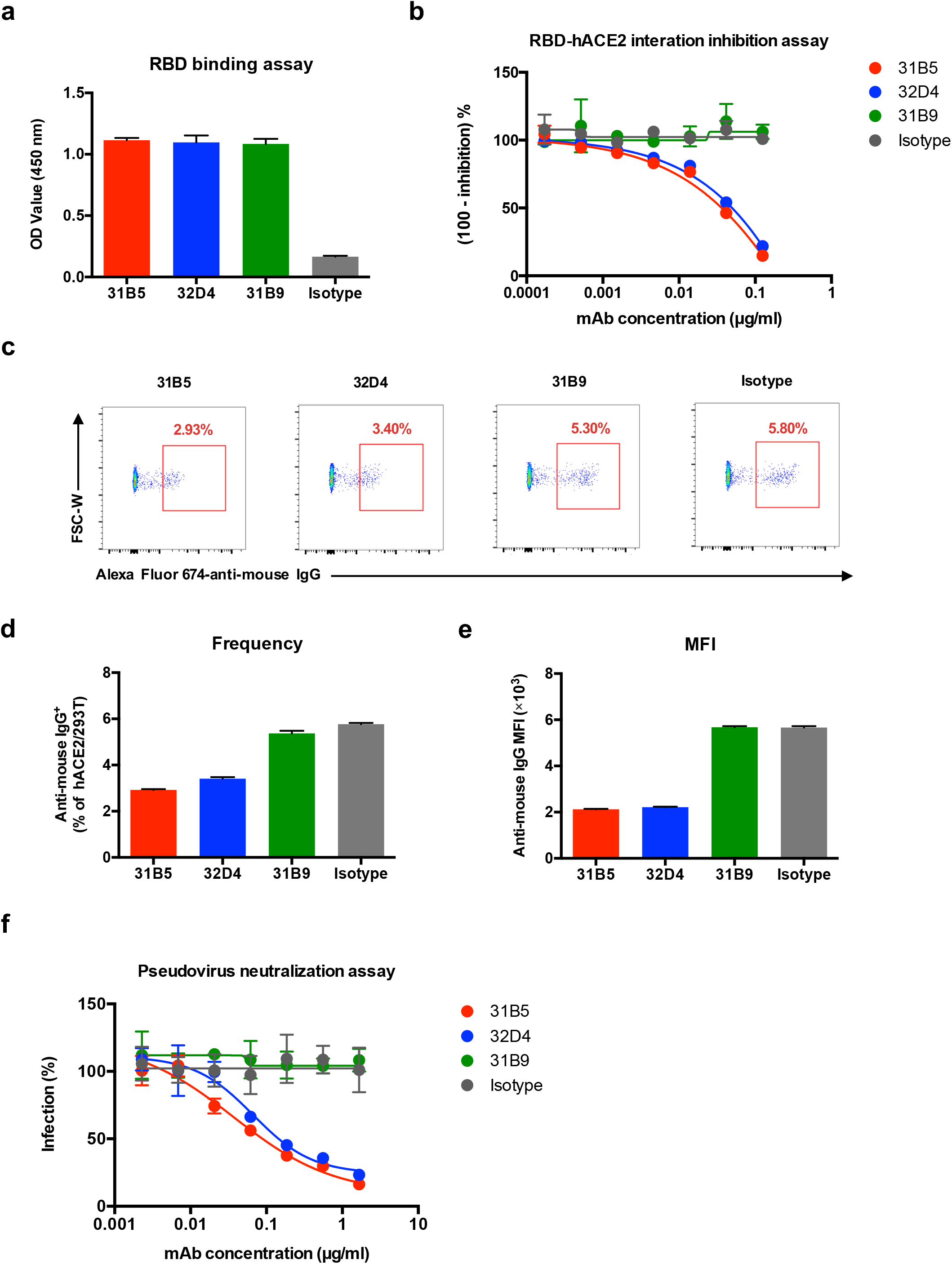
Characterization of mAbs against SARS-Cov2 RBD. **(a)** Specificity of mAbs (311mab-31B5, -32D4 and -31B9 clones) to SARS-CoV-2 RBD protein by ELISA. **(b)** ELISA analysis of SARS-CoV-2 RBD-hACE2 interaction inhibited by 311mab-31B5, -32D4 and -31B9 mAbs. **(c)** Flow cytometry analysis of SARS-CoV-2 RBD-hACE2 interaction inhibited by 311mab-31B5, -32D4 and -31B9 mAbs. The numbers adjacent to the outlined areas indicate the percentages of anti-mouse IgG^+^ hACE2-plasmid transiently transfected 293T cells, which are summarized in **(d). (e)** Mean fluorescence intensity of Alexa Fluor 647 anti-mouse IgG in anti-mouse IgG^+^ hACE2-plasmid transiently transfected 293T cells. **(f)** Antibody-mediated blocking of luciferase-encoding SARS-Cov-2 typed pseudovirus into hACE2/293T cells. The data are representative of two independent experiments with three replicates per group (a, b, d-f; error bars in a, b, d-f indicate the SD).

In conclusion, we have successfully cloned two human blocking mAbs using SARS-CoV-2 RBD-specific memory B cells isolated from recovered COVID-19 patients. These two mAbs can specifically bind to SARS-CoV-2 RBD, block the interaction between SARS-CoV-2 RBD and hACE2 receptor, and lead to efficient neutralization of SARS-CoV-2 S protein pseudotyped virus infection. Such human anti-SARS-CoV-2 RBD-hACE2 blocking mAbs hold great promise to be exploited as specific prophylactic and therapeutic agents against ongoing SARS-CoV-2 pandemic.

## Data Availability

The data in the manuscript are of availability.

## Acknowledgement

We thank Dr. Jun Huang (University of Chicago) for discussion. This work was supported by grants from the National Natural Science Fund for Distinguished Young Scholars (No. 31825011 to L.Y.) and the Chongqing Special Research Project for Novel Coronavirus Pneumonia Prevention and Control (No. cstc2020jscx-2 to L.Y.; No. cstc2020jscx-fyzx0074 to Y.C.; cstc2020jscx-fyzx0135 to Y.C.).

## Author contributions

X.C., R.L., Z.P., Q.C., Y.Y., R.Y., J.Z., L.G., Z.L, Q.H., L.X., J.T., Q.T., W.Y., L.H., and X.Y. performed the experiments. L.Y. designed the study, analyzed the data and wrote the paper with X.C., R.L., P.L., Y.C., Z.Q., X.Z., Y.W., K.D. and Z.Z.; and L.Y., Y.K. and Z.Q. supervised the study.

## Competing interests

The authors declare no competing interests.

## Materials and methods

### Human samples

The 26 COVID-19 patients enrolled in the study were provided written informed consent. Prior to blood collection, the patients were clinically recovered and tested negative qPCR for SARS-CoV-2 virus RNA. Healthy control subjects were 2 adult participants in the study. The study received IRB approval at Chongqing Public Health Medical Center (2020-023-01-KY).

### PBMC and serum collection

Blood samples were collected in cell preparation tubes with or without sodium citrate (BD Bioscience). PBMCs were isolated from blood in sodium citrate tubes using Ficoll (TBD Science), washed with PBS plus 2% FBS, suspended in cell freezing medium (90% FBS plus 10% DMSO), frozen in freezing chamber first at -80°C, and then transferred to liquid nitrogen. Sera were collected from blood without sodium citrate treatment and stored in aliquots at -80°C.

### Single-cell sorting by flow cytometry

For B cell enrichment, PBMCs were firstly stained with FITC-conjugated anti-CD19 antibody (Biolegend) on ice for 30 min. Then, FITC-CD19 stained PBMCs were enriched using anti-FITC MicroBeads (Miltenyi Biotec) by following manufacturer’s protocol. For surface staining, the enriched CD19^+^ B cells were stained with biotin-conjugated SARS-CoV-2 RBD protein (Sino Biological, 40592-V05H) at 4°C for 20 min, followed by PE-Cy7-conjugated streptavidin (eBioscience), PE-conjugated anti-CD20 antibody (Biolegend), APC-conjugated anti-human IgG (Fc) (Biolegend), APC-Cy7-conjugated anti-CD3 antibody (Biolegend), APC-Cy7-conjugated anti-CD14 antibody (Biolegend), APC-Cy7-conjugated anti-CD56 antibody (Biolegend) and APC-Cy7-conjugated LIVE/DEAD dye (Life Technologies). Cell staining was performed in PBS containing 5% mouse serum (wt/vol). For cell sorting, the stained SARS-CoV-2 RBD-specific IgG^+^ B cells were single-cell sorted into 96-well plates loaded with 10 μl catch buffer and then stored at -80°C. Catch buffer: to 1 ml of RNAase-free water (Tiangen Biotech), add 50 μl 1.5 M Tris pH 8.8 (Beijing Dingguo Changsheng Biotech) and 40 μl Rnasin (NEB).

### RT-PCR and PCR cloning

The heavy and light chain genes were PCR amplified as previously described^21^. Briefly, single-cell sorted plates were thawed on ice and added with 15 μl RT-PCR master mix following the one step RT-PCR kit protocol (Takara, RR057A) with primers for IgG VH and IgG VL. RT-PCR program: 50°C for 30 min, 94°C for 2 min, 45 cycles of 94°C for 30 sec, 57°C for 30 sec and 72°C for 1 min. Then, RT-PCR products were nested PCR-amplified with nested PCR master mix following the HS DNA polymerase kit protocol (Takara, TAK R010) with primers for IgG VH or IgG VL. Nested PCR program: 98°C for 4 min, 45 cycles of 98°C for 1 min, 57°C for 1 min and 72°C for 1 min. Next, heavy and light chain PCR products were purified and nuclease digested with Age1-HF/Sal1-HF (NEB) and Age1-HF/BsiW1-HF (NEB), respectively. The digested heavy and light chain genes were further cloned into human IgG1 heavy chain and light chain expression vectors, respectively.

### Transfection

Human embryonic kidney (HEK) 293T cells of 80-90% confluent in the 15 cm tissue culture plate were transfected with master mixture containing 9 μg heavy chain plasmid, 9 μg light chain plasmid and 60 μl *Trans*IT-293 Transfection reagent (Mirus). The culture media was changed to basal media 24 hours-post transfection. Then, the culture media was collected from the plate 2 days later.

### ELISA

50 ng of SARS-CoV-2 S1 protein (Sino Biological, 40591-V08H) or SARS-Cov2 RBD protein (Sino Biological, 40592-V08B) in 100 μl PBS per well was coated on ELISA plates overnight at 4°C. Then, the ELISA plates were blocked for 1 hour with blocking buffer (5% FBS plus 0.05% Tween 20). Next, mAbs or ten-fold diluted patient sera were added to each well in 100 μl blocking buffer for 1 hour. After washing with PBST, the bound antibodies were incubated with anti-human IgG HRP detection antibody (Bioss Biotech) for 30 min, followed by washed with PBST, then PBS and addition of TMB (Beyotime). The ELISA plates were allowed to react for 5 min and then stopped by 1 M HCl stop buffer. The optical density (OD) value was determined at 450 nm.

### ELISA-based receptor-binding inhibition assay

200 ng of hACE2 protein (Sino Biological, 10108-H08H) in 100 μl PBS per well was coated on ELISA plates overnight at 4°C. Then, the ELISA plates were blocked for 1 hour with blocking buffer (5% FBS plus 0.05% Tween 20); meanwhile, three-fold serial dilutions of mAbs or ten-fold diluted patient sera were incubated with optimal dose (based on EC_50_) of SARS-Cov2 RBD protein (Sino Biological, 40592-V05H) for 1 hour. Then, the incubated mixtures were added to ELISA plates and allowed to develop for 30 min, followed by PBST washing and anti-mouse Fc HRP antibody (Thermo Fisher Scientific). Next, the ELISA plates were washed with PBST, then PBS and added with TMB (Beyotime). After 5 min, the ELISA plates were stopped and determined at 450 nm. The half maximal inhibitory concentration (IC_50_) was determined by using 4-parameter logistic regression.

### Flow cytometry-based receptor-binding inhibition assay

311mab mAbs or isotype were incubated with optimal dose (based on EC_50_) of SARS-Cov2 RBD protein (Sino Biological, 40592-V05H) for 1 hour at RT. Then, the mixtures were incubated with 10,000 hACE2-plasmid transiently transfected 293T cells for 40 min on ice, followed by stained with Alexa Fluor 647-conjugated goat anti-mouse IgG (Biolegend) and APC-Cy7-conjugated LIVE/DEAD dye (Life Technologies).

### Pseudovirus neutralization assay

Spike protein of SARS-Cov-2 typed pseudovirus was produced as previously described^22^. Concisely, HEK-293T cells were transfected with psPAX2, pLenti-GFP and 2019-nCov S plasmids by using *Trans*IT-293 Transfection reagent (Mirus). The culture media was changed to fresh media 12 hours-post transfection. And at 64 hours after transfection, supernatants were harvested. For pseudovirus neutralization assay, three-fold serially diluted mAbs were mixed with SARS-Cov-2 typed pseudovirus for 1 hour. Then, the mixture was incubated with hACE2-expressing HEK-293T (hACE2/293T) cells overnight, followed by change of fresh media. At 40 hours-post incubation, the luciferase activity of infected hACE2/293T cells were detected by Dual-Luciferase Reporter Assay System (Promega). The percent of infection was calculated as ratio of luciferase value with mAbs to that without mAbs. The half maximal inhibitory concentration (IC_50_) was determined by using 4-parameter logistic regression.

## Notes

### Competing Interest Statement

The authors have declared no competing interest.

